# Protocol for a Real-Time Electronic Health Record Implementation of a Natural Language Processing and Deep Learning Clinical Decision Support Tool: A Use-Case for an Opioid Misuse Screener in Hospitalized Adults

**DOI:** 10.1101/2022.12.04.22282990

**Authors:** Majid Afshar, Sabrina Adelaine, Felice Resnik, Marlon P. Mundt, John Long, Margaret Leaf, Theodore Ampian, Graham J Wills, Benjamin Schnapp, Michael Chao, Randy Brown, Cara Joyce, Brihat Sharma, Dmitriy Dligach, Elizabeth S. Burnside, Jane Mahoney, Matthew M Churpek, Brian W. Patterson, Frank Liao

## Abstract

The clinical narrative in the electronic health record (EHR) carries valuable information for predictive analytics, but its free-text form is difficult to mine and analyze for clinical decision support (CDS). Large-scale clinical natural language processing (NLP) pipelines have focused on data warehouse applications for retrospective research efforts. There remains a paucity of evidence for implementing open-source NLP engines to provide interoperable and standardized CDS at the bedside. This clinical protocol describes a reproducible workflow for a cloud service to ingest, process, and store clinical notes as Health Level 7 messages from a major EHR vendor in an elastic cloud computing environment. We apply the NLP CDS infrastructure to a use-case for hospital-wide opioid misuse screening using an open-source deep learning model that leverages clinical notes mapped to standardized medical vocabularies. The resultant NLP and deep learning pipeline can process clinical notes and provide decision support to the bedside within minutes of a provider entering a note into the EHR for all hospitalized patients. The protocol includes a human-centered design and an implementation framework with a cost-effectiveness and patient outcomes analysis plan.

## INTRODUCTION

As of 2017, over 95% of hospitals in the US have adopted an electronic health record (EHR); more than 80% are collecting electronic clinical notes.^1^ Clinical decision support (CDS) and intelligent data-driven alerts are part of federal incentive programs for Meaningful Use.^2, 3^ With the increasing capacity of EHR data and financial incentives to improve quality care, hospitals are increasingly well-equipped to leverage computational resources to improve case identification and care throughput.^4^ The unstructured narrative of the electronic health record provides a rich source of information on patients’ conditions that may serve as clinical decision-support tools. Detailed medical information is routinely recorded in providers’ intake notes. Yet, this information is neither organized nor prioritized during routine care for augmented intelligence at the bedside. Moreover, clinical notes’ free text format hinders efforts to perform analytics and leverage the large domain of data. The computational methods of natural language processing (NLP) can derive meaning from clinical notes, from which machine learning algorithms can screen for conditions such as opioid misuse.

In 2020, overdose deaths from opioid misuse have soared to an all-time high with a record 93,000 deaths nationwide during the pandemic year.^5^ Substance misuse ranks second among principal diagnoses for unplanned 7-day hospital readmission rates.^6^ Screening for patients at-risk for opioid use disorders is not part of the admission routine at many hospitals, and many hospitalized patients in need are never offered opioid treatment. The high prevalence of substance use disorders in hospitalized adults exceeds rates in the general population or outpatient setting and reveals the magnitude of this lost opportunity.^7^ We previously trained a convolutional neural network (CNN) that outperformed a rule-based approach and other machine learning methods for screening opioid misuse in hospitalized patients. The CNN substance misuse classifier had greater than 80% sensitivity and specificity and demonstrated that clinical notes captured during a hospitalization may be used to screen opioid misuse.^8^

There remains a paucity of evidence on implementing clinical NLP models in an interoperable and standardized clinical decision support system for health operations and patient care.^9^ The interactions among an AI system, its users, the implementation, and the environment influence the AI interventions’ overall potential effectiveness. Few health systems have been able to accommodate the complexities of an NLP deep learning model integrated into an existing operational ecosystem and EHR.^10^ This protocol describes a cloud service designed to ingest, process, and store clinical notes as standardized and interoperable messages from a major EHR vendor in an elastic cloud computing environment. We subsequently demonstrate the use of multiple open-source tools, including an open-source NLP engine to process the EHR notes and feed them into a deep-learning algorithm for screening opioid misuse. Our resultant NLP and deep learning pipeline can process clinical notes and provide decision support to the bedside within minutes of a provider entering a note into the EHR.

To our knowledge, we offer the first protocol for a bedside implementation of an NLP-driven clinical decision support tool. We expect our protocol will serve as a guide for other health systems to leverage open-source tools across interoperable data standards and ontologies. Our goal is to describe a hospital-wide protocol and computing architecture to implement a real-time NLP-driven CDS tool. We provide an implementation framework and cost-effectiveness analysis of a tool developed for the automated screening of hospitalized adults for opioid misuse.

## METHODS

### Hospital Setting and Study Period

The NLP CDS tool will be implemented at the University of Wisconsin (UW) Hospital across the surgical and medical hospital inpatient wards. The EHR system at UW Health is Epic^©^ (Epic Systems Corporation, Verona, WI, USA). The tool is designed for hospitalized adults (18 years of age and older) and will be assessed using a pre-post quasi-experimental study design over 30 months (24 months of usual care and 6 months with implementation of automated screening). The study is a quality improvement initiative by the health system to provide an automated hospital-wide screening system for opioid misuse. The clinical study met exemption status for human subjects research by the UW Institutional Review Board.

### Pre-Intervention Period: Usual Care with Ad-Hoc Addiction Consults

UW Hospital launched an Addiction Medicine inpatient consult service in 1991 to address the high prevalence of substance use disorders in hospitalized adults. A screening, brief intervention, and referral to treatment (SBIRT) program^11^ was instituted for alcohol misuse. Screening, intervention flow sheets, and consult order sets were built into EHR-driven workflows for inpatient nurses and social workers for alcohol screening with the Alcohol Use Disorders Identification Test-Concise^12^, including a best practice alert (BPA) for patients at risk of alcohol use disorder and order sets for withdrawal treatment. For other drugs, a single screening item queries ‘marijuana or other recreational drug use,’ but no formal screening process has been in place specifically targeting opioid misuse. For patients at risk of an opioid use disorder, the current practice is ad-hoc consultations at the discretion of the primary provider.

### Post-Intervention Period: Computing Architecture and Real-Time Implementation

The technical architecture that enables the real-time, NLP CDS tool incorporates industry-leading and emerging technological capabilities. **Figure 1**. details the overall NLP CDS infrastructure to export the notes from the EHR, organize them and feed them into an NLP pipeline, input the processed text features into the opioid screener deep learning model, and deliver the resultant scores back to the bedside EHR as a BPA. The final architecture is a real-time NLP CDS tool, and the six components of the architecture are further detailed below.

**Figure 1.**
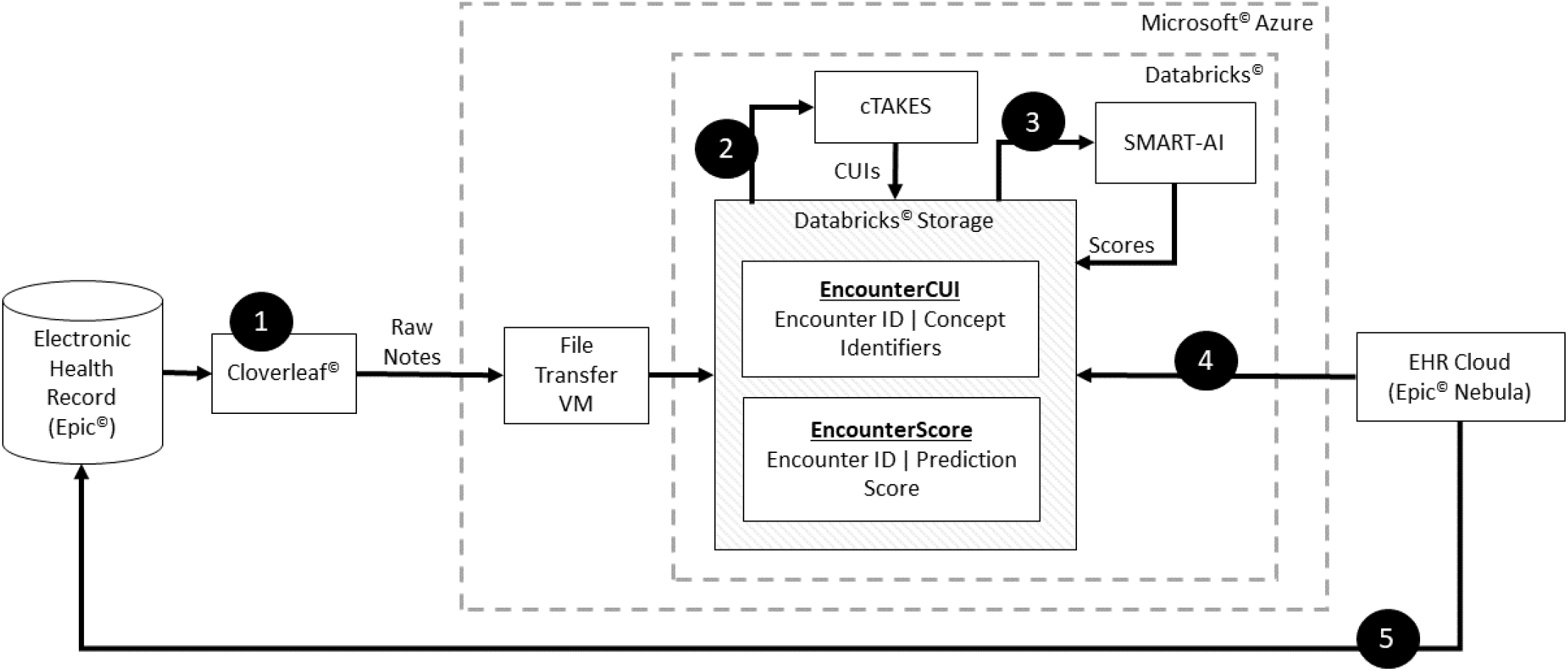
Architecture for Real-Time Natural Language Processing and Deep Learning Implementation for Clinical Decision Support Two major implementation processes are depicted. The *first process* runs a scheduled program to ingest notes from the EHR (Step 1) for each patient and organize them and relay the notes via an HL7 data feed (Cloverleaf) into the cloud computing environment and data lake (Microsoft Azure and Databricks) onto a virtual machine (Step 2). The NLP engine (cTAKES) processes the text stored on the VM and maps them to medical concepts from the National Library of Medicine’s metathesaurus (CUIs). The machine learning model receives the CUIs as inputs and stores the results in DataBricks. At regular intervals, a custom python script in Databricks does the text extraction, linguistic feature engineering via cTAKES, and storage of CUIs with appended data with patient identifiers. The CUIs serves as the input to the machine learning model (SMART-AI) at the encounter level. The output of prediction probabilities and classification are stored in a Databricks table (Step 3). The *second process* is an API call from the EHR cloud to determine if the cutpoint threshold from the machine learning model is met to trigger a best practice alert. The EHR cloud makes an HTTP call to Databricks to request the score (Step 4). The score is returned to the EHR cloud and subsequently delivered as a BPA when the provider opens the patient’s chart in the on-premise instance of the EHR at the bedside (Step 5). The full life cycle iterates every 15 minutes for a near real-time operation.

#### Component 1: Transferring Clinical Notes from EHR to Cloud Computing

HL7 refers to the standards for transferring healthcare data between data sources. Cloverleaf^©^ (Infor Cloverleaf^©^ Integration Suite) is UW’s vendor solution to serve as an Application Programming Interface (API) gateway to access the clinical narratives in the EHR via an HL7 data feed. The API extracts clinical notes from Epic^©^ and transfers them into a Microsoft Azure cloud computing environment that is under a Business Associate Agreement with UW. On-premise relays with the FTP protocol are used to transfer the clinical notes to a specified location in the Azure cloud environment.

#### Component 2: Cloud Analytic Computing Platform

In the Microsoft^©^ Azure framework (Microsoft, 2022), UW Health invoked the Databricks^©^ (Databricks Inc., San Francisco) analytic resource and services for scalable computing, data storage and querying. The open-source tools from the NLP engine and our trained, publicly available machine-learning model are hosted in Databricks. The Machine Learning model lifecycle management (MLFlow) tool in Databricks^©^ supports the data flow for the deep learning model. MLFlow creates and scores models when clinical notes are received and subsequently reports the results upon request. The final infrastructure is a scaleable and failure-resistant environment for analytic computation.

#### Component 3: Natural Language Processing Pipeline

The clinical Text Analysis and Knowledge Extraction System (cTAKES, Apache Software Foundation) builds on multiple open-source Apache projects and incorporates technologies with the Unstructured Information Management Architecture (UIMA) framework and the Apache OpenNLP natural language processing toolkit.^13^ This configuration contains several engines for sentence detection, tokenization, part-of-speech tagging, concept detection, and normalization to extract information from the clinical narrative in the EHR. cTAKES is one of the most ubiquitous NLP engines used in the clinical domain.^14^ cTAKES provides named entities from the free text that are mapped from the National Library of Medicine’s Unified Medical Language System (UMLS), which are groups of words with relevant clinical context (e.g., Drugs, Diseases, Symptoms, Anatomical Sites, and Procedures). Each named entity maps to a concept unique identifier (CUI) using the UMLS SNOMED and RxNORM ontologies. For instance, ‘heroin misuse’ from the text is assigned C0600241 as its CUI and is a separate CUI from ‘history of heroin misuse’, which is C3266350. For generalizability, we use the default cTAKES pipeline available at https://cwiki.apache.org/confluence/display/CTAKES/.

As clinical notes are entered into the EHR for an individual patient, Cloverleaf^©^ relays the notes via HL7 from Epic^©^ EHR and uses the Azure FTP server running on a virtual machine to place them in a known location within the Azure cloud environment. In 15-minute intervals, DataBricks triggers a custom python script to extract the text and feed it into the cTAKES pipeline to map and extract the CUIs. The CUIs are stored in the Azure Data Lake with appended data including patient ID, encounter ID, and note timestamp and are now ready to be fed into any machine learning model.

#### Component 4: Text Feed from NLP Pipeline into Deep Learning Model

We previously published a substance misuse screening tool using CUIs fed into a CNN called the Substance Misuse Algorithm for Referral to Treatment using Artificial Intelligence (SMART-AI).^8^

SMART-AI was trained on the first 24 hours of clinical notes from the EHR to provide enough time for robust training but lead time for the Addiction consult service to intervene before hospital discharge. For ease of implementation, the model was not trained on any specific note type and followed a timestamp approach for all notes filed within 24 hours from arrival at the hospital. Temporal validation of the classifier (trained on data between 2017 and 2019 and tested on data from 2020) at an outside hospital demonstrated an area under the precision-recall curve of 0.87 (95% CI 0.84–0.91) for screening opioid misuse. Similar results were shown in external validation at a second, independent health system.^8^ The number needed to alert to detect a true positive screen was 1.4 for opioid misuse and would create 26 alerts per 1000 hospitalized patients. This was deemed as an acceptable workload for consultation requests in live production for the UW Addiction Medicine providers. An additional retrospective review was performed at UW Health to examine sensitivity and specificity with 95% confidence intervals (CI).^15^

All notes from the first 24 hours of arrival at the UW hospital are combined into a single document per patient encounter and converted into sequences of vector representations (e.g., embeddings). The CUI embeddings define the input layer to the SMART-AI model at the encounter level. The model provides prediction probabilities for opioid misuse and stores them in a DataBricks^©^ table with a predefined cutpoint for screen positives. The SMART-AI is publicly available at https://github.com/Rush-SubstanceUse-AILab/SMART-AI

#### Component 5: Real-Time Delivery of Prediction Results

Nebula Cloud Platform is Epic^©^’s Software as a Service (SaaS) platform for integrating new technology and specifically supporting clinical prediction modeling. Nebula capabilities include the deployment of machine learning models, including a library of Epic-curated models for healthcare and custom algorithms. Our solution leverages the latter to facilitate triggers from the Epic© EHR to call out to the Databricks© environment and provide the predictions for BPAs.

In the case of SMART-AI, we designed a BPA (**Figure 2**) to trigger once a provider opens the patient’s EHR. Epic^©^ calls its Nebula component to see if a BPA should be generated. Nebula makes an HTTP call to DataBricks^©^ to request the score. The RESTful HTTP API to provide the SMART-AI model score is serviced using MLFlow. Parameters include UMLS dictionaries, model results, a flag for patient identifiers, and other necessary attributes for individual-level predictions. The score is returned to Nebula which is used to trigger a BPA if it meets the cutpoint for opioid misuse. For screen positives, the alert provides the recommendation to the clinician for consultation with UW’s Addiction Medicine consult service. The following were internal targets to meet the real-time needs of the end-user at the bedside: (1) a throughput of 1000 notes per minute (<60ms each); (2) three nines (99.9%) availability – equivalent of fewer than nine hours downtime annually; and (3) establishing an error rate threshold.

**Figure 2.**
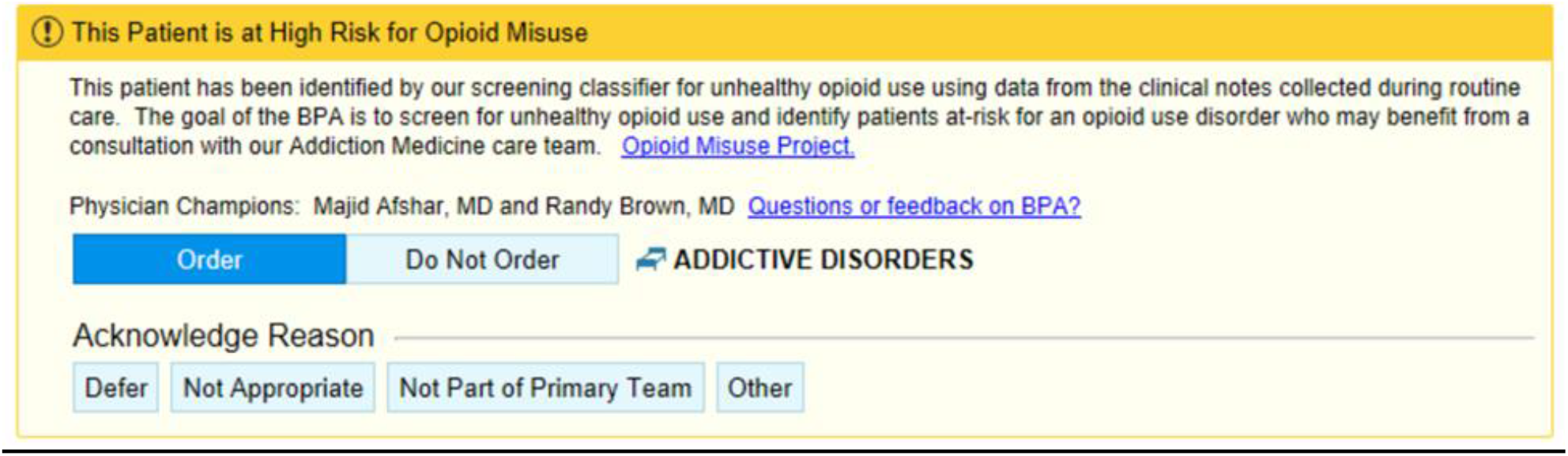
Opioid Misuse Screen Positive Clinical Decision Support with a Best Practice Alert into the Electronic Health Record (^©^2021 Epic Systems Corporation) In an iterative design with feedback from end-users, a final best practice alert (BPA) was implemented for bedside care. The BPA triggers upon opening a chart for a patient that meets the cutpoint predicted probability for opioid misuse from the NLP and Deep Learning Model (SMART-AI).

#### Component 6: Cybersecurity

Two principles of security were applied: (1) defense-in-depth and (2) zero-trust. Zero trust architecture is outlined in the National Institute of Standards and Technology publication SP 800-207.^16^ To secure access between Azure Databricks^©^ MLFlow and Epic^©^’s Nebula we employed an authentication token and IP range restriction (Databricks^©^ admin utility). The authentication token is issued via Databricks^©^ standard authentication. As a security best practice, we employed the Databricks service principal and its Databricks^©^ access token to give automated tools and systems access to Databricks^©^ resources.

### Implementation Framework

The Consolidated Framework for Implementation Research (CFIR) informed the development of the pre-implementation assessments and will be used during the rapid Plan-Do-Study-Act (PDSA) cycles.^17^ Key stakeholder interviews were planned to better understand the context and identify barriers and facilitators to implementing the BPA tool. Selected implementation strategies from the Expert Recommendations for Implementing Change (ERIC) were selected to overcome barriers.^18^ For pilot implementation, a regular cadence of meetings is planned with the implementation team to process, reflect, and evaluate barriers to implementation and use of the BPA. Process evaluation will incorporate interviews with providers and addiction specialists to understand what barriers still exist to utilizing and acting on the BPA. During pilot implementation, we will collect and summarize clinical performance data during PDSA cycles to guide clinicians and administrators to monitor, evaluate, and modify provider behavior. Using the CFIR-ERIC matching tool^19^, we will tailor relevant implementation strategies to enhance provider uptake and use of the tool. Also, during the pilot phase, we will interview providers on the hospital units beyond the pilot units to identify and explore their determinants to use of the BPA. After a pilot implementation period of three months, we will optimize provider training, enhance educational materials, and institute quality monitoring preparatory to hospital-wide rollout.

### Patient Outcomes Analysis and Power Calculation

The SMART-AI study intervention sample consists of all hospitalized patients who screen positive for opioid misuse from the NLP CDS tool. The primary effectiveness measure is the percentage of hospitalized patients in the NLP CDS intervention sample who are screened positive for opioid misuse and who received an intervention by the inpatient addiction consult service. A control sample will be derived by retrospectively applying the NLP CDS tool to all inpatient EHR records for the two years before the present study initiation in January 2023. Hospitalized patients who would have screened positive retrospectively under the NLP CDS tool will form the usual care control group.

The primary outcome is the percentage of inpatients who screened positive (or would have screened positive) based on the NLP CDS tool who receive an addiction consult with any of the following interventions: (1) receipt of opioid use intervention or motivational interviewing; (2) receipt of medication-assisted treatment (MAT); and/or (3) referral to substance use disorder treatment. The primary outcome will be reported as a percentage in the pre- and post-intervention periods and consists of substance use screening and treatment service engagement for hospitalized patients screened for opioid misuse. Secondary outcomes will include the 30-day unplanned hospital readmission rate. Criteria for unplanned hospital readmissions were adopted from the Centers for Medicare & Medicaid Services.^20^

Hypothesis testing for intervention effects will be conducted using independent tests of the difference in the proportion of patients receiving motivational interviewing, MAT, and/or referral to substance use disorder treatment. The null hypothesis is that the proportion with the primary outcome is lower (inferior) in the post-intervention period compared to pre-intervention, and the non-inferiority/equivalence margin is M, i.e., H_0_: p_1_ - p_2_ > M. The alternative one-tailed test for non-inferiority, H_1_: p_1_ - p_2_ < M, will be tested with the z-statistic.

In hospital-wide screening, we expect a prevalence of 3% of adult inpatients with opioid misuse based on prior findings of hospital-wide analyses. A total sample size of 12500 patients with 10000 in the pre-intervention 2-year period and 2500 in the post-intervention 6-month period have 85% power to detect a difference of +0.75% in the post-intervention period (3.75%) compared to the pre-intervention period (3.0%) with a non-inferiority difference of -0.5% using a one-sided z-test with significance level = 0.025.

### Cost-effectiveness analysis

Cost-effectiveness analysis estimates the incremental costs of the SMART-AI intervention (i.e., the added costs post-implementation of the SMART-AI tool in reference to usual care) and the incremental effectiveness for the primary and secondary outcomes. The health economic evaluation will determine incremental intervention costs by examining: (1) the opportunity start-up costs of implementing the SMART-AI tool; (2) the incremental medical costs resulting from usual care for hospitalized patients with opioid misuse versus SMART-AI automated screening supported care costs; and (3) the ongoing costs of administering and maintaining the SMART-AI tool.

The start-up costs of establishing SMART-AI substance use screening care will include the costs associated with developing and implementing the NLP CDS tool: (1) the cost of supporting the NLP and machine learning components and building the BPA in the EHR; and (2) training the health professionals on tool use. The incremental costs between usual care and SMART-AI automated screening supported care will be determined by calculating medical care costs before and after the implementation of the SMART-AI. Medical costs associated with the hospitalization stay and all subsequent medical costs for the 30 days following hospital admission for the pre- and post-SMART-AI intervention period will be derived from hospital billing records and presented from the single-payer (a health system) perspective.

The following three-pronged approach will be applied to identify the administration and maintenance costs associated with SMART-AI screening workflow changes introduced by the NLP CDS tool: (1) conducting in-depth interviews with hospital administrators; (2) performing activity-based observations of healthcare personnel who use SMART-AI; and (3) querying the clinician messaging system in the EHR. Average hospital compensation rates will be used for valuing healthcare personnel time costs. Research-related costs will be excluded.

### Analytical approach to cost-effectiveness analysis

Cost-effectiveness analysis is reported in terms of the incremental cost-effectiveness ratio (ICER) per additional patient who receives substance use treatment. For this study, the ICER will be calculated as the difference between pre-implementation and post-implementation intervention costs divided by the difference between pre-implementation and post-implementation intervention effectiveness as measured by rates of patient engagement with substance use treatment services (i.e., primary outcome) and 30-day hospital readmission (i.e., secondary outcome).

The usual care control group and SMART-AI intervention group will each be characterized by the pathway probabilities of substance use treatment receipt and meeting the primary outcome. The pathway probabilities of patients’ engagement with inpatient substance use consult, brief intervention/motivational interviewing (MI), and referral to substance use treatment for both study groups will result in 8 treatment combinations based on (Addiction consult/MI/MAT/Referral x pre-/post-implementation). The differential costs pre- and post-SMART-AI intervention will be determined as the difference in the weighted sum of the individual pathway costs, using the pathway probabilities as weights for the intervention and control groups. Effectiveness will be determined as the difference in rates of hospitalized patients engaging with substance use disorder treatment pre- and post-implementation of the SMART-AI automated screening aided care for the intervention and control groups. The ICER will be calculated as follows:

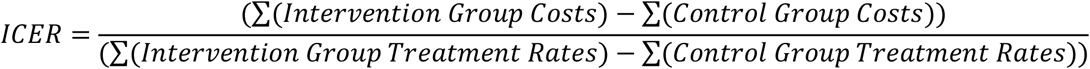

Sensitivity analyses will introduce uncertainty in substance use treatment receipt rates and costs for the intervention and control groups. Monte Carlo-based simulation estimation will use the rates of substance use treatment service uptake observed in the intervention and control groups as a reference to simulate a cohort of post-implementation hospitalized patients and a cohort of usual care hospitalized patients. The ICER per additional individual who receives an inpatient substance use consult, brief intervention, MI, MAT, or referral to substance use treatment will be calculated by drawing a random sample with replacement from the observed distributions for health-care costs (μ_COSTi_) and substance use treatment services (μ_TRTi_) for intervention and control groups. This process will then be repeated (N=1000) to produce bootstrap estimates of the 95% confidence intervals for the ICER per additional individual who receives an inpatient substance use consult, brief intervention, MI, MAT, or referral to substance use treatment. These probabilistic sensitivity analyses will estimate the elasticity of the differential cost per patient relative to differential substance use treatment service rates for intervention and control groups.

## RESULTS

Early-stage investigations were performed to assess the AI system’s predictive performance in a retrospective setting and evaluate the human factors surrounding the BPA before initiating the quasi-experimental clinical study. During the retrospective review of SMART-AI at UW Health, a random sample of 100 adult patient encounters (with an over-sampling of patients with ICD codes for substance use) in 2021 was extracted and reviewed by an inpatient physician and clinical informatics expert. SMART-AI performed similarly to prior published reports for screening opioid misuse with a sensitivity of 93% (95% CI 66%-99%) and specificity of 92% (95% CI 84%-96%).

Before the deployment of SMART-AI, approvals were received across hospital committees for inpatient operations, EHR super users, clinical decision support, and nursing documentation. The proposal protocol was also reviewed by the Center for Clinical Knowledge Management to confirm no competing interests or roles with existing protocols for screening substance use conditions existed in the health system. In addition, SMART-AI was reviewed by UW’s Clinical AI and Predictive Analytics Committee. A model review form providing details on the clinical problem, model value proposition, model description, proposed workflow integration, internal validation, and monitoring strategy (including fairness and equity) was reviewed and approved by a multidisciplinary committee of clinicians, informaticians, bioethicists, executive leadership, and data scientists. The planned workflow from introduction to implementation is depicted in **Figure 3**.

**Figure 3.**
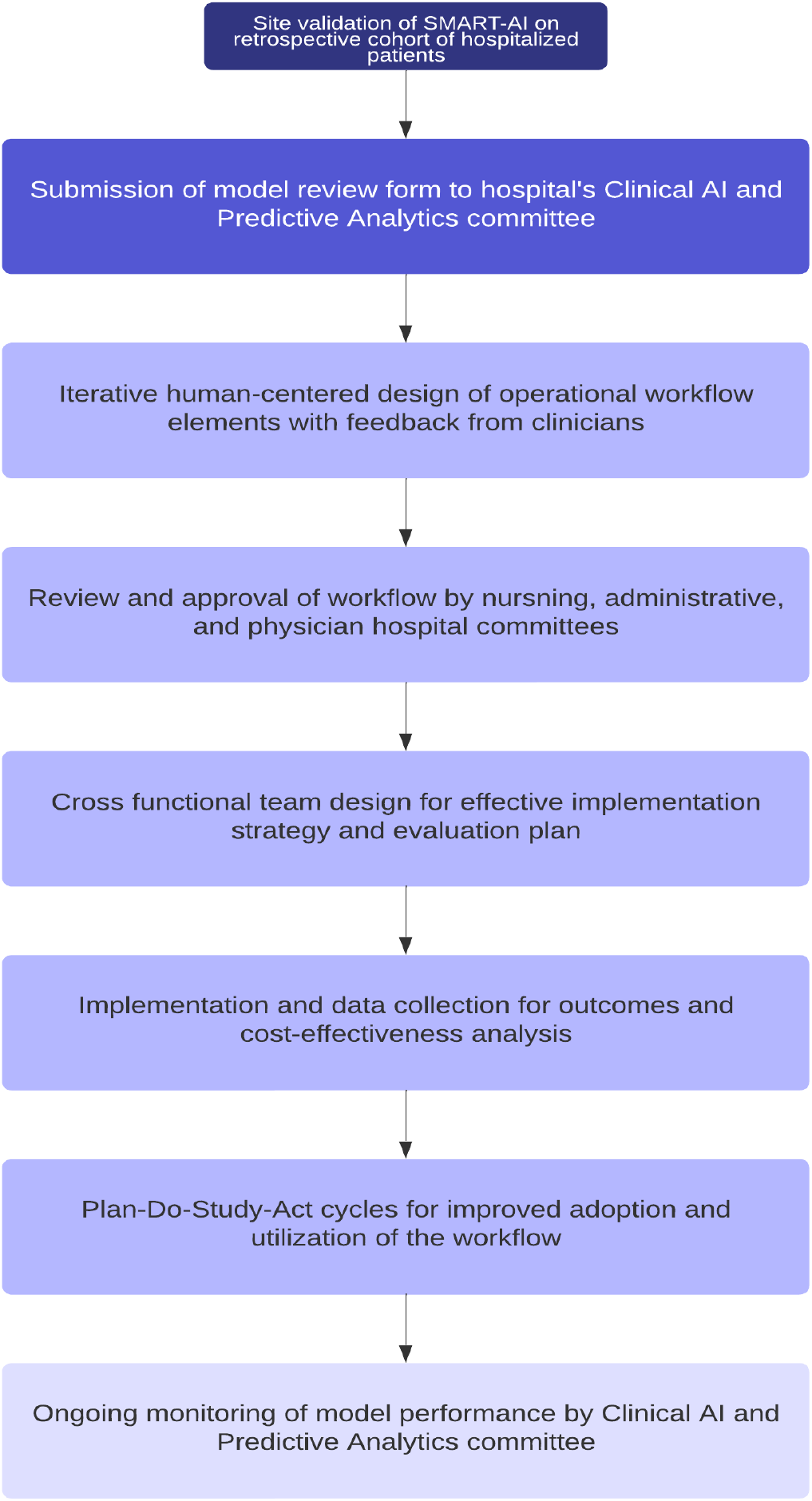
Flow Diagram for the Process to Bedside Implementation and Evaluation

An end-user interview guide and survey were developed to examine the user acceptability of the BPA. Open-ended questions were asked about barriers and facilitators to using the BPA. Five interviews were conducted (three nurse practitioners, one family medicine resident, and one surgical attending), and the responses led the production team to create an educational flyer, modify the BPA with more details and options for consultation refusal, and modify when and where the BPA would trigger. **Figure 2** is the final production version of the BPA for deployment. Dissemination efforts included Grand Round presentations to the Addiction Medicine Division, Department of Family Medicine, and notification via the hospital’s weekly electronic newsletter.

The longest delay in operational workflow and architecture was receiving cybersecurity approvals, especially with data exchange of protected health information between the Microsoft^©^ and Epic^©^ cloud vendors. An additional six months of delays occurred to achieve acceptable security monitors and checks. The Go-live of SMART-AI into the EHR was scheduled for January 2023.

## DISCUSSION

The digital era in medicine continues to grow exponentially in both the quantity of unstructured data collected in the EHR and the number of prediction models developed for detection, diagnostic, prognostic, and therapeutic guidance. In parallel, the clinical NLP field has grown in its capabilities with the advent of transformer architectures and more affordable and efficient cognitive computing of big data.^21^ However, a major bottleneck remains in the successful implementation of NLP and deep learning models into clinical practice. Much of the progress in NLP has focused on information retrieval and extraction^22^ but the application of these methods at scale with the combination of software developers and operations (DevOps) remains challenging at healthcare institutions. We offer one of the first protocols that detail the components for a real-time NLP-driven CDS system for healthcare delivery at the bedside. We further detail an implementation framework with human-centered design principles and a planned iterative process to evaluate the cost-effectiveness and health outcomes to improve screening in opioid misuse.^23^

Applied clinical NLP has predominantly remained a rule-based approach but statistical machine-learning models are now the leading method in the research literature.^14^ Few vendors that provide NLP services rely entirely on machine learning and a gap remains in effectively applying NLP models into bedside EHRs that go beyond disease detection with explicit, keyword mentions.^4, 24^ Several barriers exist with neural language models, including the need to remove PHI so the trained models may be shared and the computational requirements to run complex deep learning models in a production environment.^25^ We offer solutions for both approaches using a feature engineering approach to map free text to coded vocabulary and describe a large computing infrastructure with a connection between a data science cloud platform and the electronic health record to support direct data feeds into any machine learning model. The NLP CDS pipeline accomplishes efficiencies in data standardization and scalability^26^ for successful implementation and is extensible to other NLP engines. The benefit of augmented intelligence remains unknown and is the next step with our healthcare outcomes and cost-effectiveness analysis in a clinical study.

Our implementation framework is largely guided by a team of implementation scientists, supported by the university’s Clinical and Translational Science Award (CTSA). We leveraged our CTSA’s Dissemination and Implementation (D&I) Launchpad to help bridge the gap between evidence-based research and practice.^27^ The D&I Launchpad works to accelerate the pace of disseminating research findings and increase the adoption and implementation of effective interventions, leading to sustainable practice and policy changes. They employ strategies from implementation science, design thinking, and human-centered engineering for better integration of AI technologies in health systems. As part of the pre-implementation phase, we assessed contextual factors that may impact implementation by engaging both adopters, who are the decision-makers, and end-users, who are the main implementers, of the tool.^28^ We performed qualitative interviews with end-users to evaluate the need for the tool and the BPA design. We involved adopters early in the process to inform the intervention/implementation process through consultations during the design, feasibility testing, and implementation phases. An iterative process ensued to address constraints and contextual factors that affect adoption and implementation in our health system.

During pre-implementation, the project team clarified roles with project management with an assessment for the readiness of the clinical workflow approved through hospital committee meetings and individual interviews with end-users. Our health system is an early adopter of AI governance with a review process similar to other health systems.^29, 30^ The Clinical AI and Predictive Analytics committee follows the minimum information about clinical artificial intelligence modeling (MI-CLAIM Checklist).^31^ The offline validation of our model incorporated principles from multiple reporting guidelines on prediction models, bias and fairness, and validation.^31, 32^ Clinical evaluation after the go-live of SMART-AI will follow the reporting guideline for the early-stage clinical evaluation of decision support systems driven by artificial intelligence (DECIDE-AI).^33^ During implementation, reviews by the Clinical AI and Predictive Analytics committee will include quarterly evaluations for the sustained effectiveness of the tool, audit its fairness across parity groups, and examine for alert fatigue.

The build of an enterprise-wide AI infrastructure for data-driven CDS is an important feature of a data-driven Learning Health System (LHS). At UW, LHS activities dating back to 2013 established an evidence-based framework with a series of organizational-level quality improvement (QI) interventions.^34^ In 2020, UW Health reaffirmed its strategic plan embedding discovery and innovation as well as diversity, equity, and inclusion in clinical care. Successful implementation included coaching staff and administrative leaders to work in PDSA with lean management to get the problem, analysis, corrective actions, and action plan down on a single sheet of large (A3) paper, also known as “A3” thinking.^35^ A rapid PDSA cycle is important in the advent of AI-driven interventions that require rigorous evaluation for implementation or de-implementation.

The deployment of medical AI systems in routine clinical care presents an important yet unfulfilled opportunity^36^, and our protocol aims to close the gap in the implementation of AI-driven CDS.^37^ Our protocol implementation for an enterprise-wide production environment of an AI opioid misuse screener provides a model for other health systems to use to bring NLP models into practice for CDS. We highlight opportunities to leverage the expertise of our Applied Data Science team to employ the open-source tools for feature engineering and model development inside a larger infrastructure with vendor support of hardware and software dependencies. Given the sensitive nature of healthcare data, the biggest challenge remains to ensure high standards for cybersecurity and meet the privacy requirements for protecting patient data.

## Data Availability

The raw EHR data are available upon request due to ethical and legal restrictions imposed by the University of Wisconsin-Madison Institutional Review Board. The original data derives from the institutions EHR and contains patients protected health information (PHI). Data are available from University of Wisconsin Health Systems for researchers who meet the criteria for access to confidential data and have a data usage agreement with the health system.

https://github.com/Rush-SubstanceUse-AILab/SMART-AI

https://cwiki.apache.org/confluence/display/CTAKES/

## DATA AVAILABILITY

The raw EHR data are available upon request due to ethical and legal restrictions imposed by the University of Wisconsin-Madison Institutional Review Board. The original data derives from the institution’s EHR and contains patients’ protected health information (PHI). Data are available from the University of Wisconsin Health Systems for researchers who meet the criteria for access to confidential data and have a data usage agreement with the health system. Only the final trained model that is fully de-identified with a vocabulary of mapped concept unique identifiers is open-source and available at: https://github.com/Rush-SubstanceUse-AILab/SMART-AI. Our de-identification approach has been previously described.^38^

## ACKNOWLEDGEMENTS

The authors acknowledge support from the University of Wisconsin Institute for Clinical and Translational Research supported by the Clinical and Translational Science Award (CTSA) program, through the NIH National Center for Advancing Translational Sciences (NCATS) grant (2UL1TR002373). Research was also supported by the National Institute on Drug Abuse of the National Institutes of Health (NIDA R01DA051464; CJ, DD, MO, MA, RB, BS), the National Library of Medicine THYME project (NLM R01LM010090; DD), and the National Institute of Diabetes and Digestive and Kidney Diseases (NIDDK R01DK126933). The content is solely the responsibility of the authors and does not necessarily represent the official views of the NIH or the other funding sources listed above.

## AUTHOR CONTRIBUTIONS

Majid Afshar, Frank Liao, Brian Patterson, Sabrina Adelaine, Felice Resnik, Cara Joyce, Marlon Mundt, Dmitriy Dligach, and Matthew Churpek led the conception and design of the study and provided study supervision. Authors Afshar, Liao, Mundt, Ampian, Wills, Chao had full access to all the data in the study and take responsibility for the integrity of the data and the accuracy of the data analysis. Cara Joyce, Brihat Sharma, and Dmitriy Dligach could not access the original data directly because of limitations in the data use agreement but take responsibility for the accuracy of the data analysis and did have access to all the data presented in the manuscript. Authors Afshar, Adelaine, Mundt, Leaf, Ampian, Wills, Schnapp, Chao, Joyce, Sharma, Burnside, Resnik, Mahoney, Dligach, Patterson, Churpek, and Liao performed the analysis and/or interpretation of data. Administrative, technical and material support were provided by Authors Afshar, Patterson, Adelaine, Long, Ampian, Wills, Schnapp, and Liao. All authors reviewed the manuscript and provided edits and revisions. All authors take responsibility for the integrity of the work as a whole, from inception to the finished article, and all authors approved the final version submitted. Dr. Afshar was responsible for the decision to submit the manuscript.

